# Inflammatory and neurodegenerative serum protein biomarkers increase sensitivity to detect disease activity in multiple sclerosis

**DOI:** 10.1101/2023.06.28.23291157

**Authors:** Tanuja Chitnis, Ferhan Qureshi, Victor M. Gehman, Michael Becich, Riley Bove, Bruce A. C. Cree, Refujia Gomez, Stephen L. Hauser, Roland G. Henry, Amal Katrib, Hrishikesh Lokhande, Anu Paul, Stacy J. Caillier, Adam Santaniello, Neda Sattarnezhad, Shrishti Saxena, Howard Weiner, Hajime Yano, Sergio E. Baranzini

## Abstract

**Background/Objectives:** Serum proteomic analysis of deeply-phenotyped samples, biological pathway modeling and network analysis were performed to elucidate the inflammatory and neurodegenerative processes of multiple sclerosis (MS) and identify sensitive biomarkers of MS disease activity (DA).

**Methods:** Over 1100 serum proteins were evaluated in >600 samples from three MS cohorts to identify biomarkers of clinical and radiographic (gadolinium-enhancing lesions) new MS DA. Protein levels were analyzed and associated with presence of gadolinium-enhancing lesions, clinical relapse status (CRS), and annualized relapse rate (ARR) to create a custom assay panel.

**Results:** Twenty proteins were associated with increased clinical and radiographic MS DA. Serum neurofilament light chain (NfL) showed the strongest univariate correlation with radiographic and clinical DA measures. Multivariate modeling significantly outperformed univariate NfL to predict gadolinium lesion activity, CRS and ARR.

**Discussion:** These findings provide insight regarding correlations between inflammatory and neurodegenerative biomarkers and clinical and radiographic MS DA.

**Funding:** Octave Bioscience, Inc (Menlo Park, CA).

## INTRODUCTION

Multiple sclerosis (MS) is a chronic inflammatory demyelinating disease of the central nervous system, with a variable presentation and heterogenous disease course^1, 2^. While the exact pathophysiology of MS remains elusive, inflammatory and degenerative processes are believed to play a role^3–5^. Identifying disease-specific biomarker sets may assist in predicting diverse disease courses, classifying patients to high versus low risk for disease activity (DA) and progression (DP) and may also provide insights into mechanisms of new inflammatory DA^6, 7^. Multivariate models reflecting multiple biological pathways involved in the complex pathophysiology of MS will likely increase the predictive accuracy of these biomarkers^6^.

Most studies have focused primarily on serum neurofilament light chain (sNfL) as a liquid biomarker in MS. Levels of sNfL have been associated with neurodegeneration in MS and correlate with measurements of DA, including the presence of gadolinium-enhancing (Gd+) lesions and clinical relapse^6–11^. For example, a recent study of over 500 sNfL samples, a significant elevation in sNfL was observed after a clinical relapse only when associated with a Gd+ lesion^12^. In the 3 months after a Gd+ lesion, an average 35% elevation in sNfL (p<0.0001) was reported versus samples from patients in remission^12^. Similarly, an average 32.3% elevation in sNfL was observed at the time of, or prior to a Gd+ lesion (p=0.002) versus remission^12^. However, this individual biomarker is limited in sensitivity and specificity and is only marginally elevated during relapse^12^.

The MS relapse disease process includes both an inciting inflammatory peripheral and central immune activation, with subsequent central nervous system damage in the form of myelin and neuronal degradation^1^, as evidenced by immunological studies, and response to specific immune cell–targeting disease-modifying therapies. Moreover, MS genetic susceptibility studies demonstrate involvement of T and B cell–associated genes.^13^ Thus, including additional inflammatory and neurodegenerative protein biomarkers, can offer deeper insights and reveal stronger correlations to clinical and radiographic DA than sNfL alone.

In the search for more specific and comprehensive sets of markers, several cytokines, chemokines, and other immune-related molecules have been associated with DA in patients with MS. For example, baseline levels of cerebrospinal fluid (CSF) proteins CXCL13, CXCL12, IFN*γ*, TNF, sCD163, LIGHT, and APRIL have been associated with evident DA compared with no evidence of DA (NEDA) in patients with MS^14^. CSF levels of glial biomarkers, including glial fibrillary acidic protein (GFAP) and Chitinase 3-like-1 (CHI3L1 or YKL-40), have been associated not only with DA but also with disability^15, 16^. Masvekar et al. found that the additive model of IL12p40 and CHI3L1 correlated with new MS lesion activity^17^. MS disease severity has been associated with alterations in proteins reflecting astrocyte (MMP7, SERPINA3, GZMA, and CLIC1) and microglial activation (DSG2 and TNFRSF25)^18^.

To provide new insights and identify sensitive biomarkers of new MS DA, both radiographic (new Gd+ lesions) and clinical (relapses), we evaluated over 1100 serum proteins in over 600 samples from three MS cohorts. Biological pathway modeling and network analysis were performed to ensure comprehensive representation of MS neurophysiology and to gain insights into the inflammatory, immune and neurodegenerative process in MS.

## RESULTS

Results from this study are divided into the following three sections: protein down-selection analysis, univariate analysis of each endpoint outlined in the Methods, and multivariate modeling of each endpoint.

### Custom assay panel (CAP) proteins

The final list of CAP proteins can be found in Table 1. GFAP was not one of the original 1411 proteins described in the Methods section. It was added to our panel after much of the development work described in this report was completed because of its strong association with several DA- and DP-related endpoints. Therefore, it was not part of the analysis. The remaining 20 proteins in Table 1were carried forward.

**Table 1.** | Custom assay panel (CAP) proteins picked in the down-selection process.

| **Analyte** | **Protein name/alias** | **UniProt ID** |
| --- | --- | --- |
| APLP1 | Amyloid $\beta$ Precursor-Like Protein 1 | P51693 |
| CCL20 | C-C Motif Chemokine Ligand 20 (MIP-3) | P78556 |
| CD6 | Cluster of Differentiation 6 | P30203 |
| CDCP1 | CUB Domain Containing Protein 1 | Q9H5V8 |
| CNTN2 | Contactin 2 | Q02246 |
| COL4A1 | Collagen-α1(IV) Chain | Q07325 |
| CXCL9 | Chemokine (C-X-C Motif) Ligand 9<br>(MIG: Monokine Induced by $\gamma$ Interferon) | O43927 |
| CXCL13 | C-X-C Motif Chemokine Ligand 13, BLC | P02462 |
| FLRT2 | Fibronectin Leucine-rich Repeat Transmembrane | O43155 |
| GFAP | Glial Fibrillary Acidic Protein | P14136 |
| GH | Growth Hormone, Somatotropin | P01241 |
| IL-12B | Interleukin-12 Subunit Beta | P29460 |
| MOG | Myelin Oligodendrocyte Glycoprotein | Q16653 |
| NEFL | Neurofilament Light Polypeptide Chain, NfL | P07196 |
| OPG | Osteoprogenin, TNFRSF11B | O00300 |
| OPN | Osteopontin | P10451 |
| PRTG | Protogenin | Q2VWP7 |
| SERPINA9 | Serpin Family A Member 9 | Q86WD7 |
| TNFRSF10A | Tumor Necrosis Factor Receptor Superfamily<br>Member 10A (TRAIL-R1), DR5, Death Receptor 5 | O00220 |
| TNFSF13B | Tumor necrosis factor superfamily member 13B,<br>B Cell Activating Factor, BAFF | Q9Y275 |
| VCAN | Versican Core Protein, Versican Proteoglycan | P13611 |

### Univariate results

As a precursor to a formal univariate analysis of each endpoint, we grouped samples by label and represented the results as box plots to look for qualitative trends in concentration for each class at a population level. Gd lesion samples were separated into groups for zero, one, two, and three or more Gd+ lesions (Fig. 1). Purely binary endpoints (clinical relapse status [CRS] and low versus high annualized relapse rate [ARR]) were split according to positive and negative labels (Fig. 2).

**Fig. 1.**
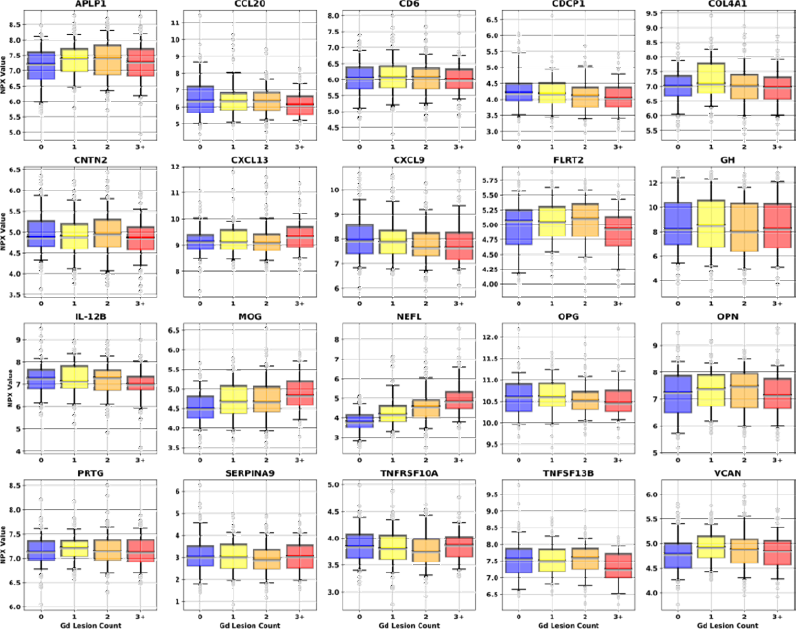
|Univariate dependence of the 20 CAP protein concentrations on Gd-enhanced lesion count. Different color boxes correspond to the lesion count in that population of samples (blue for zero lesions, yellow for one, orange for two, and red for three or more). The black line through each box shows the median (50^th^ percentile) of the population. The height of each box shows the interquartile range (25^th^-75^th^ percentile). The whiskers show the central 90% of the distribution (5^th^-95^th^ percentile). The 5% of outliers furthest from the median are drawn as open light gray circles. CAP custom assay panel; Gd gadolinium; NPX normalized protein expression.

**Fig. 2.**
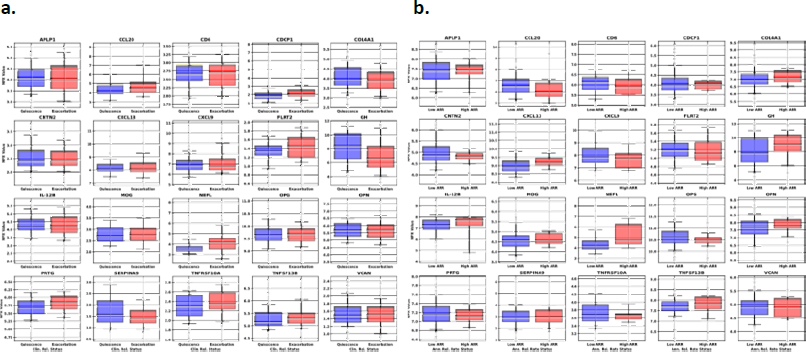
| Univariate box plots for the CAP proteins separation of samples(a) and box plots of the univariate separation of CAP proteins for low and high ARR(b) a: Univariate box plots for the CAP proteins separation of samples taken during “quiescence” (remission, blue boxes) or “exacerbation” (relapse, red boxes). b: Box plots of the univariate separation of CAP proteins for low and high ARR. The box/whisker ranges and outliers are plotted analogously to Fig. 1. ARR annualized relapse rate; CAP custom assay panel.

To quantitatively investigate the univariate significance, Pearson’s correlation and Student’s *t* test were used. The former checked for linear correlation between protein concentration and lesion count (clipped at five lesions). The latter checked the binary endpoints, univariate separation of samples associated with no Gd lesions from those associated with ≥1, as well as CRS and ARR status. Pearson’s R and Student’s *t* were computed for each protein in Fig. 3 to assess the relationship between protein concentration and all endpoints.

**Fig. 3.**
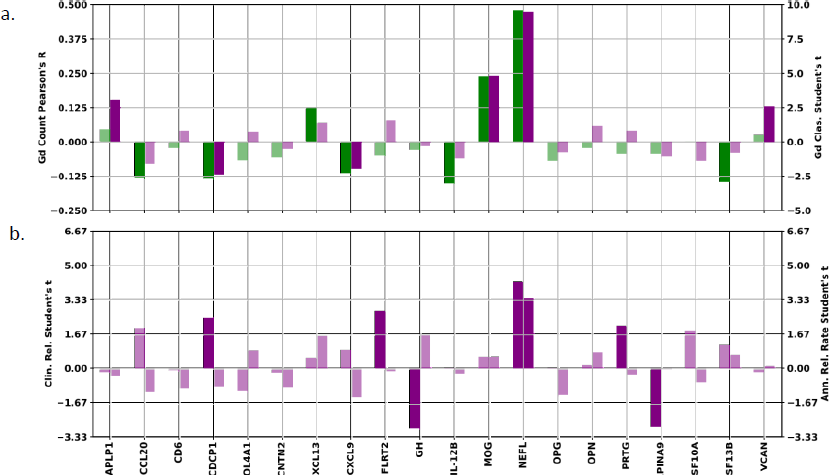
| Univariate statistical tests for all endpoints. a: Pearson’s R correlation between NPX concentration and Gd lesion count capped at five (green bars, left axis) and Student’s *t* statistic for separation of samples associated with zero lesions from those with one or more by NPX concentration (purple bars, right axis) for each protein. b: Student’s *t* statistic for separation of samples associated with clinically inactive from those with clinically active disease state (left axis) and those associated with low from high ARR (right axis) by NPX concentration. Bars corresponding to statistical tests showing a p-value >0.05 have been drawn in a lighter shade of the same color to denote their lack of statistical significance. ARR annualized relapse rate; Gd gadolinium; NPX normalized protein expression.

Examination of the bar charts in Fig. 3 allowed us to check the directionality and significance of all univariate statistical tests. Of particular interest were proteins showing consistency between the direction of the correlation/separation for all endpoints. Agreement between correlation and separation was particularly important for Gd lesion count and presence, since these two endpoints are not independent of each other. Proteins showing a test statistic with opposite signs between the two Gd tests did not pass the significance threshold. Furthermore, only CDCP1 shows a difference between the Gd endpoints and the other two, with negative correlation/separation for Gd lesions, but positive mean shift with clinically active disease state (CDCP1 shows no significant relationship to our ARR endpoint).

The proteins passing the significance threshold in each of the univariate analyses were (with test statistics and p-values):

- Gd lesion regression: NfL (2.8×10^−29^), MOG (7.8×10^−8^), IL-12B (8.0×10^−4^), TNFSF13B (1.1×10^−3^), CDCP1 (2.7×10^−3^), CCL20 (3.5×10^−3^), CXCL13 (5.4×10^−3^), CXCL9 (1.1×10^−2^)
- Gd lesion classification: NfL (1.0×10^−19^), MOG (1.9×10^−6^), APLP1 (2.3×10^−3^), VCAN (9.9×10^−3^), CDCP1 (1.8×10^−2^), CXCL9 (4.7×10^−2^)
- CRS: NfL (5.0×10^−5^), GH (4.4×10^−3^), SERPINA9 (5.3×10^−3^), FLRT2 (6.6×10^−3^), CDCP1 (1.6×10^−2^), PRTG (4.2×10^−2^)
- ARR: NfL (7.9×10^−4^)

### Multivariate results

Forward selection curves, regression, and classification Gd lesion analysis, as well as the classification of CRS and AAR are shown in Fig. 4.

**Fig. 4.**
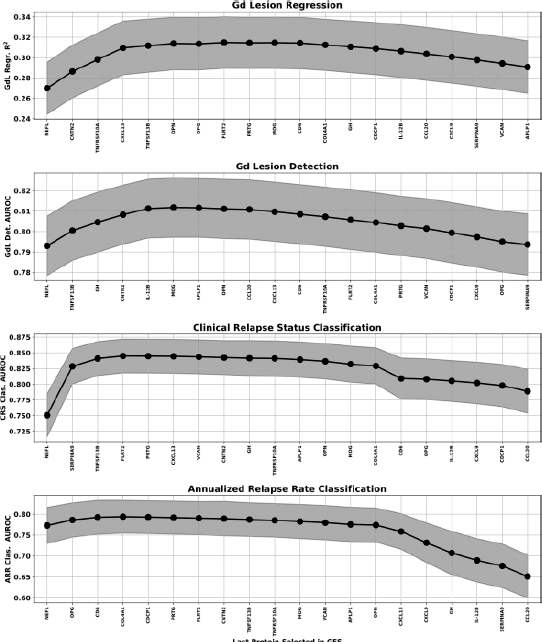
| GFS curve for the regression (top) and classification (bottom three) analysis of all endpoints. The protein features selected for each of the multivariate analyses were: The points represent the mean over the bootstrap splits and the shaded region represents the standard deviation.

- Gd lesion regression: NfL, CNTN2, TNFRSF10A, CXCL13, TNFSF13B
- Gd lesion classification: NfL, TNFSF13B, GH, CNTN2, IL-12B
- Clinical relapse status: NfL, SERPINA9, TNFSF13B, FLRT2
- Annualized relapse rate: NfL, OPG, CD6 Note that the regression analysis was clipped at a lesion count of five, as it was for the univariate correlation analysis. AUROC area under the receiver operator characteristic; ARR annualized relapse rate; CRS clinical relapse status; Gd gadolinium; GFS greedy forward selection; NfL neurofilament light chain.

Multivariate model performance for all four analyses, along with permutation-based feature importance were plotted in Fig. 5. Results for the Gd lesion regression analysis were presented as a two-dimensional histogram depicting the distribution of actual versus predicted lesion count for the regression analysis, and as receiver operating characteristic (ROC) curves for the classification analysis. The central line in each ROC band represented the mean of the ROC across all bootstraps; band width was the uncertainty in that mean. The regression analysis panel of Fig. 5 also included the best fit line through the results drawn in solid black with the root mean square error as a shaded gray region. “Perfect agreement” (actual = predicted) was drawn for reference in dashed gray. For the classification analysis panels, we included the following two feature sets in addition to the greedy forward selection (GFS) proteins for each endpoint: NfL only, and all features except NfL. Feature importance (was estimated by permuting the values of each protein individually and checking the performance decrease of the model for each bootstrap built on the GFS features).

**Fig. 5.**
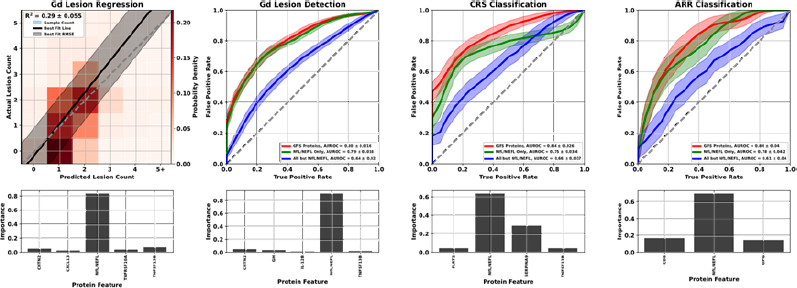
| Multivariate modeling results for all four analyses: Gd lesion count estimation (zero through five or more, left panel), Gd lesion detection (center left panel), CRS classification (center right panel), and ARR classification (right panel) Each analysis was plotted with the performance plot above a feature importance bar graph for the GFS proteins. ARR annualized relapse rate; CRS clinical relapse status; Gd gadolinium; GFS greedy forward selection.

We tabulated the results of all four multivariate analyses in Table 2. For the Gd lesion detection endpoint, we quantified model performance in the following three different ways:

- General DA (GDA): separation of samples with no Gd lesions from those with any positive count of lesions
- Subtle DA (SDA): separation of samples with no Gd lesions from those with only one lesion
- Extreme DA (EDA): separation of samples with no Gd lesions from those with three or more lesions

**Table 2.** | Classification and regression performance for all analyses of all endpoints for different feature sets^a^.

| Analysis | GFS proteins | NfL/NEFL only | All except NfL/NEFL |
| --- | --- | --- | --- |
| Gd lesion count regression (R<sup>2</sup>) | 0.290±0.056 | 0.270±0.056 | 0.105±0.036 |
| GDA (AUROC) | 0.801±0.032 | 0.792±0.032 | 0.636±0.040 |
| SDA (AUROC) | 0.695±0.050 | 0.694±0.048 | 0.598±0.052 |
| EDA (AUROC) | 0.917±0.032 | 0.908±0.032 | 0.700±0.058 |
| Clinical relapse status (AUROC) | 0.847±0.054 | 0.754±0.070 | 0.659±0.078 |
| Annualized relapse rate status (AUROC) | 0.797±0.082 | 0.776±0.088 | 0.544±0.128 |

<sup>a</sup>We report mean performance and its uncertainty (standard deviation) across all bootstrap simulations.
AUROC, area under the receiver operating characteristics curve; EDA extreme disease activity; Gd gadolinium; GDA general disease activity; GFS genome-based peptide fingerprint scanning; NfL neurofilament light chain; SDA subtle disease activity.

While we only trained lesion detection models on GDA, we included model performance metrics for the SDA and EDA endpoints as well in our discussion of Gd lesion detection. Additionally, we checked Gd lesion regression performance on models constructed from both NfL only and all proteins except NfL for comparison to the classification endpoint.

Comparison of the actual versus estimated Gd lesion count (left panel, Fig. 5) showed that the regression model slightly under-predicts Gd lesion count for samples with no lesions and over-predicts it for those with five or more, while the model’s accuracy for lesion counts of 1-2 anchored the performance of the model. While regression models performed acceptably, results from the classification analysis were considerably stronger. For all Gd lesion analyses, NfL had the largest effect (Table 3; Fig. 5). NfL performance as the strongest biomarker was further reflected in the feature importance plots at the two left bottom panels of Fig. 5. However, there is an incremental improvement in performance between NfL only and the GFS proteins, particularly in the Gd lesion regression analysis. Furthermore, the models using all proteins except NfL were significantly better than random chance. There is clearly signaling for this endpoint in the other proteins that is drowned out by the performance of NfL for predicting Gd lesions.

**Table 3.**
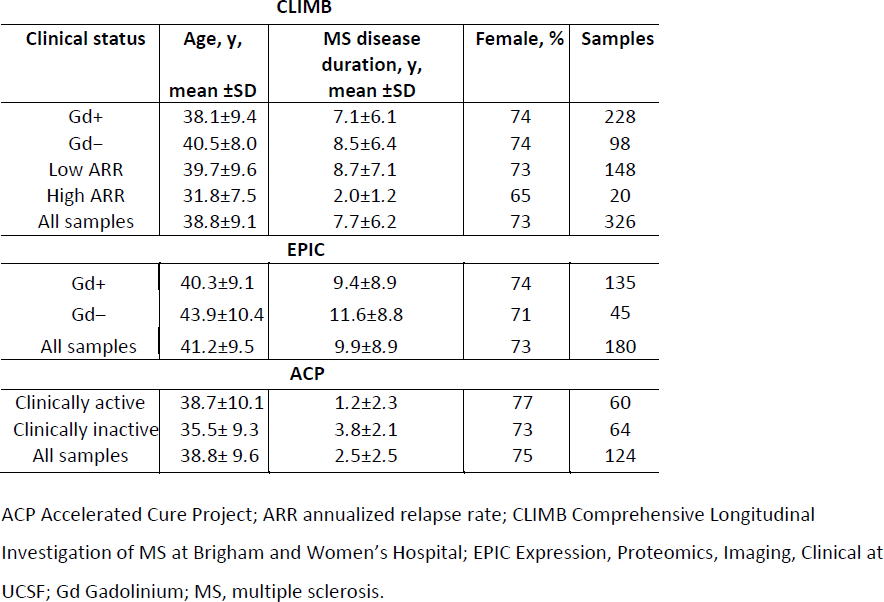
| Demographic and clinical breakdown of all three patient cohorts CLIMB.

While NfL was the strongest performer for CRS and ARR as well, it’s lead over the other proteins was considerably attenuated. GFS protein performance for CRS outperformed all endpoints except EDA, and achieved similar ARR results to that of GDA. Multivariate separation of samples by CRS and ARR was significantly better than chance and significantly better than univariate NfL. Consistent with the Gd lesion analysis, multivariate models trained on all features except NfL displayed an area under the receiver operator characteristic (AUROC) significantly better than random chance.

### Biological interpretation

The biological context of the 21 proteins listed in Table 1 was analyzed in the following two ways: direct spatial, functional, and gene expression correlations to a curated set of open-source databases, and expanded graph techniques using the Scalable Precision Medicine Open Knowledge Engine (SPOKE).

For the direct correlation analysis, protein concentration values were associated with data from the Human Protein Atlas^19^ (aggregated lymphoid tissue, peripheral blood immune cell type, and brain region proteomic data) as well as the Allen Brain Atlas^20^ (brain structure, cell type, and transcriptomic data). Protein-protein interaction modeling was performed by inputting proteins into STRING^21^ for network construction. Physically and functionally associated proteins that exhibited a minimum interaction score of 0.7 (“high confidence”) were classified as interacting with each CAP protein. Markov Clustering^22, 23^ was used to detect distinct subgraphs of interconnected proteins. Topological surveillance and centrality metric calculations were performed in Cytoscape^24^. Enrichr^25^ was leveraged to functionally annotate protein subgraphs.

Comparative proteomic analysis across human organs, tissues, and cell types helped home in on MS-relevant information by isolating organ-specific blood analytes to address the blood’s pervasive nature. This revealed the MS-specific locality of CAP proteins, facilitating downstream directionality assessment and revealing a rich repertoire of cell types related to MS. Constitutive expression of these proteins helped to facilitate downstream mechanistic modeling. The resultant mapping of the 21 CAP proteins onto 10 biological hallmarks of MS is presented in the left panel of Fig. 6. The 10 hallmarks were each subdivided into two to three related biological processes. CAP proteins were then sorted into these processes based on their correlations with the databases and tools mentioned.

**Fig. 6.**
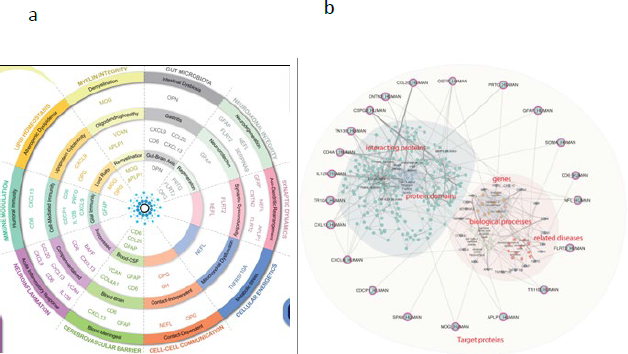
| CAP proteins sorted into biological processes and grouped into 10 MS hallmarks(a) and SPOKE graph visualization of biological neighborhood of CAP proteins(b) a: CAP proteins sorted into biological processes and grouped into 10 MS hallmarks using each protein’s correlation to spatial, functional, and gene expression data. b: SPOKE graph visualization of biological neighborhood of CAP proteins. Using each CAP protein as input results in a fully connected module including encoding genes, directly interacting proteins (and their domains), biological processes and a short list of related diseases. CAP custom assay panel; MS multiple sclerosis; SPOKE Scalable Precision Medicine Open Knowledge Engine.

The SPOKE Neighborhood Explorer (https://spoke.rbvi.ucsf.edu) allows targeted exploration of any component of the SPOKE knowledge graph. SPOKE is a comprehensive graph with millions of biomedical concepts and has been previously utilized for drug repurposing^26^, to conduct genetic analyses^27^, and for clinical predictions^28^. SPOKE has integrated data from close to 40 databases, including ChEMBL, OMIM, LINCS, and the Human Protein Atlas, among others. As such, SPOKE contains all human genes and proteins, more than a million pharmaceutical compounds, more than 7000 diseases, and a comprehensive representation of signaling and metabolic pathways. Further, biological interpretation of the 21 proteins also included spatial expression profiling, Protein-Protein interaction modeling, and Gene Set Enrichment. The SPOKE knowledge graph for the CAP proteins is shown in the right panel of Fig. 6.

## DISCUSSION/CONCLUSIONS

We evaluated multivariate analyses of blood serum biomarkers from three independent cohorts and identified twenty proteins that were strongly associated with increased clinical and radiographic activity of MS. To quantify MS activity, we focused principally on the presence of Gd+ lesions compared with patients lacking such lesions. We also examined the following two clinical measures of MS relapse activity: clinician assessment of relapse state at or near the time of blood draw, and the ARRs in the time leading up to the draw.

Feature down-selection processes can inherently introduce bias. To minimize bias, we deliberately attempted to balance weight and normalization strategies. For example:

1. Weighting each feature importance by the AUROC of the model in which it appeared. When AUROC is calculated on the same data on which the model was trained, it tends to bias the result in favor of proteins that perform well in large models. Larger models are more prone to overfitting noise in the training data, which will lead to systematically higher AUROC values.
2. Normalizing the vector of feature importance values for each model to 1 tends to favor proteins that perform better in models with only a few features. Since the total importance score for models with fewer features is split among fewer proteins compared with ones with larger feature sets, the values themselves will be numerically higher.

There are two points that strongly favor multivariate modeling over univariate NfL regarding the Gd endpoint. First, the optimized multivariate model significantly outperformed univariate NfL in every framing of the problem of using serum protein chemistry to predict Gd lesion activity. Second, we find statistically significant performance in a multivariate modeling even when the model ignores NfL concentrations. Both statements hold for an endpoint that should be most advantageous to the performance of univariate NfL (Gd lesions), and the difference was even larger for the clinical relapse endpoints. Furthermore, univariate NfL lacks specificity, as NfL levels are known to be elevated for neurodegenerative conditions other than MS^29^.

NfL showed the strongest univariate correlation with the radiographic and clinical measures of DA examined in this study. Furthermore, multivariate techniques showed increased performance compared with NfL alone for all analyses of all endpoints. Similarly, multivariate models without NfL were still significantly more performant than chance for all endpoints, suggesting signaling from the other CAP proteins as well. The inclusion of additional inflammatory and neurodegenerative protein biomarkers can provide deeper insights and reveal stronger correlations to clinical and radiographic DA than NfL individually. Cytokines, chemokines, and other immune-related molecules have consistently been associated with DA in patients with MS and they constitute an attractive target for interrogation in biological samples from patients at different stages of their disease course. Additionally, measuring protein concentration has several advantages over transcriptional profiling, including higher stability and more straightforward biological interpretation of the results. A biological pathway–centered approach using a subset of those shown in the left panel of Fig. 6 is likely to be a successful strategy for planning future investigation.

In addition to overall efficacy of the models used to examine the endpoints in this study, information from a broader panel of serum biomarkers allowed novel insights into the pathophysiology of new Gd lesions and clinical relapse. This includes the identification of chemokines that are significantly altered by DA mediated by inflammation or immune response. A multi-protein panel like the one developed in this study has the capability to capture the state of a patient’s MS from multiple angles, allowing for a fuller picture of their pathophysiology.

This study leveraged three large, well-characterized cohorts of patients with MS, and evaluated well over 1000 protein biomarkers using highly sensitive assays and applied network modeling to the findings to provide new insights into MS. We used a systems biology framework to contextualize the mechanism-of-action of selected serum protein biomarkers with respect to MS DA. Through a complementary integration of machine learning and functional network analysis, we were able to shed light on the heterogeneous pathophysiological underpinnings of MS and unveil the orchestrated crosstalk between the various molecular facets of the disease. The analysis presented here was, however, purely cross sectional. Large-scale longitudinal studies will be necessary to better understand MS and its evolution over time to unlock the potential of personalized MS care.

The end goal of the research program, the beginning of which is presented in this article, is a MS DA test that is fully validated in a clinical trial. Such a test would have tremendous clinical utility for many issues in MS care:

1. *Identification of active relapse*: MS relapses can be quite subtle, especially early in the disease course, and can be easily confused with recrudescence of symptoms in the setting of stressors (“pseudo-flares”) or conditions other than MS. This can lead to either the misattribution of an unrelated symptom to an MS relapse or to an early relapse being misattributed to some other clinical event. An inexpensive, clinically simple, precise, repeatable test for MS relapses would go a long way toward reducing both types of errors. Response to relapses often includes an escalation in disease-modifying therapy (DMT) or steroid doses, both of which can have side effects.
2. *Prediction of impending relapse*: A test like the one we describe here has the potential to also serve as a leading indicator of impending relapses ahead of clinical presentation. This capability would need to be tested and validated clinically to quantify the scale of such a lead time.
3. *Confirmation of NEDA status*: NEDA is the clinical gold standard for MS care. A NEDA designation indicates the patient’s clinical and radiographic activity are held in check. A test like this one could alert a patient’s MS care team to otherwise subclinical (or sub-radiographic) DA.
4. *Assessment of patient-specific longitudinal changes relative to previous tests*: An underlying truth beneath many scientific discoveries is that differential measurements tend to be simpler and more accurate than absolute ones. This is likely true of quantitative DA measurements as well. Some absolute level of “DA score” would be instructive to both patient and clinician, but deviations from baseline levels for an individual patient would be more so.
5. *Response to DMTs*: The ability to quantify the level of DA with a test like the one proposed could serve as an endpoint for clinical trials of DMT drugs and help clinicians to evaluate the efficacy of a DMT for a particular patient in less time than it would take for clinical or radiographic evidence to present. This would greatly enhance the quality of life and increase the “health span” of patients with MS and offer insight into patient adherence to their treatment plan.

Analytical validation of this DA panel will be followed by clinical validation studies to verify association with DA endpoints (primarily Gd lesions) in multiple independent cohorts. Expansion of the test’s clinical utility will be investigated with future studies to evaluate biomarker correlations with endpoints associated with MS DP, therapy selection, and differential diagnosis.

## METHODS

### Materials

Serum samples from three deeply phenotyped cohorts were analyzed for protein levels and associated with clinical and radiographic endpoints to select features for inclusion in a custom assay panel. The three endpoints in this study were as follows:

1. Presence of Gd lesions for samples were defined as samples for which the blood draw was performed within 30 days of a contrast-enhancing magnetic resonance imaging, with the count of Gd lesions determined by a neuroradiologist.
2. Clinically active MS versus clinically inactive MS samples were defined as samples for which the blood draw was performed during a state of active relapse or inactive remission as defined by a clinician.
3. High versus low ARR samples were defined as those for which the blood draw was performed, and corresponding ARR was determined (high: ≥1.0 and low: ≤0.2 relapses per year).

These three endpoints were taken from three different cohorts of patients and samples as follows:

1. Comprehensive Longitudinal Investigation of MS at Brigham and Women’s Hospital (CLIMB). CLIMB Endpoints: radiographically defined relapse status using Gd lesions (primary), ARR (secondary).
2. Expression, Proteomics, Imaging, Clinical at UCSF (EPIC). EPIC Endpoint: radiographically defined relapse status using Gd lesions.
3. Accelerated Cure Project (ACP). ACP Endpoint: Clinically Defined Relapse Status - active versus inactive.

This study did not include human subjects but included human serum samples obtained from three MS centers under informed consent of all participants. A summary of the salient demographic and clinical features from each cohort is presented in Table 3.

### Ethics statement

The study protocol and study procedures were approved by institutional review boards and independent ethics committees at each study site. The University of California San Francisco Institutional Review Board granted ethical approval for the EPIC Cohort. Massachusetts General Brigham Human Research Committee granted ethical approval for the CLIMB Cohort. Western Institutional Review Board, Copernicus Group IRB, Sheperd Center Research Review Committee, Institutional Review Board for Human Research at St. Joseph’s Hospital and Medical Center, University of Massachusetts Medical School Committee for the Protection of Human Subjects in Research, Ohio State University Biomedical Institutional Review Board, Beth Israel Deaconess Medical Center Committee on Clinical Investigations, John Hopkins medicine Office of Human Subjects Research Institutional Review Board, and the Southwestern Medical Center Institutional Review Board all granted ethical approval for the ACP Cohort.

### Analytical methods

For the purposes of the screening studies, up to 1411 proteins were measured using two separate immunoassay platforms. The first panel of 1196 proteins were analyzed using Proximity Extension Assay technology on the Olink™ Platform^30^. Protein concentrations were reported as NPX values (normalized protein expression), which provide expression levels relative to the other samples included on the plate and within the batch. An additional panel of 215 proteins were analyzed using xMAP® technology immunoassays at Myriad RBM, Inc. (RBM). Absolute protein concentrations from the RBM platform were determined using calibrated standard curves.

Exploratory data analysis was conducted to filter noise, reduce dimensionality, and avoid collinearity. Univariate significance was combined with multivariate importance from models generated with randomly selected combinations of different numbers of proteins to select features for inclusion into the custom assay panel. Biomarkers selected as features in the panel were investigated for relevance and interactions using biological network models. A 21-plex custom assay panel was then manufactured and analytically validated^31^ to establish the following specifications and parameters: accuracy, precision, sensitivity, specificity, reference ranges, stability (reagents and samples), diurnal variation, drug interference, and assay robustness. The custom assay panel has been manufactured to include calibrators to report results in absolute concentration and a fit-for-purpose analytical validation has been performed. All samples previously run in the biomarker screening studies were reanalyzed using the custom assay panel.

### Biostatistical methods

The program outlined in this report can be organized into the following two phases: the down-selection of the final 21 protein analytes and the studies that use those proteins as features to examine the three study endpoints. The former balanced information about all three endpoints to select a final ensemble of proteins for use as features in a more detailed analysis of all three endpoints in the latter phase. All analyses were performed in the python^32^ programming language, making use of the SciPy^33^ and Scikit-Learn^34^ packages for statistical tests and machine learning models respectively.

### Protein down selection

The protein down-selection phase of this analysis proceeded along two parallel tracks to identify the features most strongly associated with all three study endpoints. First, we looked at the univariate correlation between each of 1411 proteins as follows: 1196 on the Olink platform and 215 from RBM. Gd lesions were treated as a binary variable used to classify samples based on their presence or absence. This allowed us to treat this endpoint consistently with the other two, which are inherently binary in nature. We computed the AUROC for separating the positive from negative samples for all three endpoints for all 1411 proteins and ranked them in decreasing order for each, paying special attention to analytes that showed a strong association with more than one.

To avoid biasing our multivariate feature selection process toward proteins that performed well in one but not another model architecture (e.g. tree-based vs. linear models), we investigated several of the following model types: logistic regression, support vector classifiers, and random forest classifiers. Furthermore, we did not have an *a priori* estimate of the optimal number of features in each model, so we tested models with 3-21 proteins in steps of three. Because of its strong univariate performance in our data and its well-established association with many aspects of MS pathophysiology in the published literature, we used NfL (NfL is referred to by its gene name “NEFL” in the Olink platform, so we used the two names interchangeably in the analysis of these data) as the seed for multivariate feature selection analyses. We then randomly selected proteins in addition to NfL to fill out the set number of features (3-21, by threes) and used them to construct models of all three architecture types. This was repeated 100,000 times for each combination of model size and architecture. All models were trained on the entire dataset and evaluated for efficacy against the endpoint under consideration by calculating AUROC. After generating this ensemble of models, we extracted feature importance values from each one and tracked the feature importance for each protein over all the models in which it appeared. The following importance values for features from each model architecture were taken to be:

1. In logistic regression models, feature importance was taken to be the variance-normalized absolute value of each feature coefficient. Protein concentration was expressed in NPX for this study, which is analogous to the log of the absolute concentration of each protein. The variance across all samples therefore did not vary as widely across protein as it would if we were using linear concentration values.
2. In support vector classifiers, we took feature importance to be the feature coefficient absolute value. We used a linear kernel for our support vector classifier) models so that feature coefficients were well defined.
3. In random forest classifier models, we simply used the Gini coefficients attached to the model.

Feature importance vectors were normalized to unit sum for each model. When averaging the feature importance value for each protein across all models in which it appeared, we weighted each importance by the AUROC of its model so that proteins appearing in highly efficacious models would be favored over ones in models with poor performance. We then ranked each protein by its average AUROC-weighted feature importance so that we could quantify the multivariate performance of each protein separately from its univariate performance. We selected the top performers across each of our six lists.

### Application of selected proteins to study endpoints

After selecting the top 20 performing proteins (20, not 21 because one spot in the panel was saved for a desired protein being added to the Olink platform [see Results for further discussion]), we next evaluated their performance in the clinically relevant endpoints. The Gd lesion endpoint was investigated using both a classification and a regression approach (i.e., different models were trained to both classify each serum sample as being associated with some number of Gd lesions or not, and to estimate the number of Gd lesions associated with that sample). Investigation of the CRS involved the analysis of blood serum taken from patients during a clinically active relapse (exacerbation) or during a period of clinical stability, or inactive status (quiescence), following an approach similar to that taken for lesion presence classification. Finally, we addressed the ARR endpoint by considering the question of whether each sample had ARR <0.2 relapses per year (low) or >1 relapse per year (high). Recognizing that the modest sample size and low positivity rate of ARR (6.9%, or 13 out of 188 samples) impact the power of this analysis, the same overall strategy as the two previous binary endpoints was used.

Due to the finite timescale on which brain lesions will be enhanced under Gd contrast, we precleaned our Gd lesion data by discarding all serum samples drawn more than 30 days from (before or after) the magnetic resonance imaging from which the lesion count was extracted. We accounted for batch-to-batch variability in relative quantitation by measuring bridge normalization samples across all assays.

The univariate analysis of all three endpoints was performed using the bridge-normalized NPX values described above. The multivariate analysis for each endpoint used those same NPX values after correction for the following clinical variables: age, sex, and MS disease duration. An ordinary least squares linear regression model was fitted to the batch-normalized NPX values of the Gd-negative samples for each protein using the clinical variables as features. This estimate was then subtracted from all sample NPX values to make the demographic/clinical correction. The residual from this correction process was used as features in multivariate modeling.

Throughout each stage of the multivariate modeling process, an ensemble of bootstrap simulations was generated to control for overfitting. The data for each endpoint was randomly split into train (two-thirds) and test (one-third) subsets 1000 times. A different model of the same configuration was then trained on each bootstrap split, and model performance was taken to be the mean across the ensemble (with uncertainty parameterized by the square root of the variance). We chose bootstrap simulations instead of k-fold cross validation because the particular random split in the cross validation could have small effects on the outcome of some of the modeling efforts. The large number of random splits generated in the bootstrap process smoothed the contribution from the small number of outlier samples in our data.

Both the regression and classification analyses were performed on an optimized subset of the entire panel chosen using GFS of protein features, with Pearson’s correlation coefficient (R^2^) and AUROC, respectively, as the target metric and average over the ensemble of bootstrap splits. GFS starts with the top-performing univariate protein then checks each of the remaining features to construct the most performant two-protein model. With that in hand, each of the remaining proteins is checked in turn to see which combination provides the best three-protein performance. The process continues until all available proteins are included in the model. Optimal model size is then determined as the feature set size where performance is no longer significantly improved by adding an additional protein feature. In general, this function reaches a plateau value of optimal performance and then turns over as extra features are added that contribute only noise to the model. This is often a global maximum value of all possible combinations of features but was taken for the purposes of our study as the optimal model size and performance.

## DATA AVAILABILITY

Access to data can be provided after a research proposal is submitted to the corresponding author and a data sharing agreement is in place.

## Supporting information

STARD Checklist

## ACKNOWLEDGMENTS

Editorial assistance was provided by Maureen Wallace-Nadolski, PhD, of The Lockwood Group (Stamford, CT) and was supported by funding from Octave Bioscience, Inc (Menlo Park, CA).

## AUTHOR CONTRIBUTIONS

- Study concept and design: T.C., F.Q., V.G., M.B., R.B., B.C., R.G., S.H., R.H., A.K., H.L., A.P., S.C., A.S., N.S., S.S., H.W., H.Y., S.B.
- Data acquisition, analysis, or interpretation: T.C., F.Q., V.G., M.B., R.B., B.C., R.G., S.H., R.H., A.K., H.L., A.P., S.C., A.S., N.S., S.S., H.W., H.Y., S.B.
- Statistical analysis: V.G., M.B., A.K., H.L., S.B.
- Drafting of the manuscript: T.C., F.Q., V.G., M.B., S.B.
- Critical review of the manuscript: T.C., F.Q., V.G., S.B.
- Approval of the final article: T.C., F.Q., V.G., M.B., R.B., B.C., R.G., S.H., R.H., A.K., H.L., A.P., S.C., A.S., N.S., S.S., H.W., H.Y., S.B.

## REFERENCES

1. Baecher-Allan, C., Kaskow, B. J., & Weiner, H. L. Multiple sclerosis: mechanisms and immunotherapy. Neuron 97, 742–768 (2018).

2. Reich, D. S., Lucchinetti, C. F., & Calabresi, P. A. Multiple sclerosis. N Engl J Med 378, 169–180 (2018).

3. Chitnis, T., & Weiner, H. L. CNS inflammation and neurodegeneration. J Clin Invest 127, 3577–3587 (2017).

4. Macaron, G., & Ontaneda, D. Diagnosis and management of progressive multiple sclerosis. Biomedicines 7, 56 (2019).

5. Wingerchuk, D. M., & Carter, J. L. Multiple sclerosis: current and emerging disease-modifying therapies and treatment strategies. Mayo Clin Proc 89, 225–240 (2014).

6. Barro, C., et al. Serum neurofilament as a predictor of disease worsening and brain and spinal cord atrophy in multiple sclerosis. Brain 141, 2382–2391 (2018).

7. Comabella, M., & Montalban, X. Body fluid biomarkers in multiple sclerosis. Lancet Neurol 13, 113–126 (2014).

8. Disanto, G., et al. Serum neurofilament light: A biomarker of neuronal damage in multiple sclerosis. Ann Neurol 81, 857–870 (2017).

9. Mellergård, J., et al. Cerebrospinal fluid levels of neurofilament and tau correlate with brain atrophy in natalizumab-treated multiple sclerosis. Eur J Neurol 24, 112–121 (2017).

10. Novakova, L., et al. Monitoring disease activity in multiple sclerosis using serum neurofilament light protein. Neurology 89, 2230–2237 (2017).

11. Varhaug, K. N., et al. Neurofilament light chain predicts disease activity in relapsing-remitting MS. Neurol Neuroimmunol Neuroinflamm 5, e422 (2018).

12. Rosso, M., et al. Temporal association of sNfL and gad-enhancing lesions in multiple sclerosis. Ann Clin Transl Neurol 7, 945–955 (2020).

13. Sawcer, S., et al. Genetic risk and a primary role for cell-mediated immune mechanisms in multiple sclerosis. Nature 476, 214–219 (2011).

14. Magliozzi, R., et al. The CSF profile linked to cortical damage predicts multiple sclerosis activity. Ann Neurol 88, 562–573 (2020).

15. Comabella, M., et al. Cerebrospinal fluid chitinase 3-like 1 levels are associated with conversion to multiple sclerosis. Brain 133, 1082–1093 (2010).

16. Modvig, S., et al. Cerebrospinal fluid levels of chitinase 3-like 1 and neurofilament light chain predict multiple sclerosis development and disability after optic neuritis. Mult Scler 21, 1761–1770 (2015).

17. Masvekar, R., Phillips, J., Komori, M., Wu, T., & Bielekova, B. Cerebrospinal fluid biomarkers of myeloid and glial cell activation are correlated with multiple sclerosis lesional inflammatory activity. Front Neurosci 15, 649876 (2021).

18. Masvekar, R., Wu, T., Kosa, P., Barbour, C., Fossati, V., & Bielekova, B. Cerebrospinal fluid biomarkers link toxic astrogliosis and microglial activation to multiple sclerosis severity. Mult Scler Relat Disord 28, 34–43 (2019).

19. Karlsson, M., et al. A single-cell type transcriptomics map of human tissues. Sci Adv 7, (2021).

20. Allen Institute for Brain Science. Allen Cell Types Database—Human MorphologyElectrophysiology. celltypes.brain-map.org/data. RRID:SCR 014806 (2015).

21. Szklarczyk, D., et al. STRING v11: protein-protein association networks with increased coverage, supporting functional discovery in genome-wide experimental datasets. Nucleic Acids Res 47, D607-d613 (2019).

22. Enright, A. J., Van Dongen, S., & Ouzounis, C. A. An efficient algorithm for large-scale detection of protein families. Nucleic Acids Res 30, 1575–1584 (2002).

23. van Dongen, S. Graph clustering by flow simulation. PhD thesis, Center for Math and Computer Science (CWI), (2000).

24. Shannon, P., et al. Cytoscape: a software environment for integrated models of biomolecular interaction networks. Genome Res 13, 2498–2504 (2003).

25. Kuleshov, M. V., et al. Enrichr: a comprehensive gene set enrichment analysis web server 2016 update. Nucleic Acids Res 44, W90–97 (2016).

26. Himmelstein, D. S., et al. Systematic integration of biomedical knowledge prioritizes drugs for repurposing. Elife 6, (2017).

27. Himmelstein, D. S., & Baranzini, S. E. Heterogeneous network edge prediction: a data integration approach to prioritize disease-associated genes. PLoS Comput Biol 11, e1004259 (2015).

28. Nelson, C. A., Bove, R., Butte, A. J., & Baranzini, S. E. Embedding electronic health records onto a knowledge network recognizes prodromal features of multiple sclerosis and predicts diagnosis. Journal of the American Medical Informatics Association 29, 424–434 (2021).

29. Varhaug, K. N., Torkildsen, Ø., Myhr, K. M., & Vedeler, C. A. Neurofilament light chain as a biomarker in multiple sclerosis. Front Neurol 10, 338 (2019).

30. Slenter, D. N., et al. WikiPathways: a multifaceted pathway database bridging metabolomics to other omics research. Nucleic Acids Res 46, D661–D667 (2018).

31. Qureshi, F., et al. Analytical validation of a multi-protein, serum-based assay for disease activity assessments in multiple sclerosis. Proteomics Clin Appl, e2200018 (2023).

32. Van Rossum, G., & Drake, F. L. Python 3 *Reference Manual* (2009).

33. Virtanen, P., et al. SciPy 1.0: fundamental algorithms for scientific computing in Python. Nat Methods 17, 261–272 (2020).

34. Pedregosa, F., et al. Scikit-learn: machine learning in Python. J Mach Learn Res 12, 2825–2830 (2011).

